# Major discrepancy between factual antibiotic resistance and consumption in South of France: analysis of 539,037 bacterial strains

**DOI:** 10.1101/2020.02.10.19016188

**Authors:** Ousmane Oumou Diallo, Sophie Alexandra Baron, Gregory Dubourg, Hervé Chaudet, Philippe Halfon, Sabine Camiade, Béatrice Comte, Stéphanie Joubert, Arnaud François, Philippe Seyral, François Parisot, Jean-Paul Casalta, Raymond Ruimy, Christophe Maruejouls, Jean-Christophe Achiardy, Sophie Burignat, Joseph Carvajal, Edouard Delaunay, Sandra Meyer, Pierre-Yves Levy, Patricia Roussellier, Patrick Brunet, Claude Bosi, Philippe Stolidi, Jean-Pierre Arzouni, Gisele Gay, Pierre Hance, Philippe Colson, Didier Raoult, Jean-Marc Rolain

## Abstract

**Introduction:** The burden of antibiotic resistance is currently estimated by mathematical modeling, without real count of resistance to key antibiotics. Here we report the real rate of resistance to key antibiotics in bacteria isolated from humans during a 5 years period in a large area in southeast in France.

**Methods:** We conducted a retrospective study on antibiotic susceptibility of 539,107 clinical strains isolated from hospital and private laboratories in south of France area from January 2014 to January 2019. The resistance rate to key antibiotics as well as the proportion of bacteria classified as Difficult-to-Treat (DTR) were determined and compared with the Mann-Whitney U test, the χ2 test or the Fisher’s exact test.

**Results:** Among 539,037 isolates, we did not observe any significant increase or decrease in resistance to key antibiotics for 5 years, (oxacillin resistance in *Staphylococcus aureus*, carbapenem resistance in enterobacteria and *Pseudomonas aeruginosa* and 3^rd^ generation cephalosporin resistance in *Escherichia coli* and *Klebsiella pneumoniae*). However, we observed a significant decrease in imipenem resistance for *Acinetobacter baumannii* from 2014 to 2018 (24.19% to 12.27%; p=0.005) and a significant increase of ceftriaxone resistance in *Klebsiella pneumoniae* (9.9% to 24,03%; p=0.001) and *Enterobacter cloacae* (24,05% to 42,05%; p=0.004). Of these 539,037 isolates, 1,604 (0.3%) had a DTR phenotype.

**Conclusion:** Over a 5-year period, we did not observe a burden of AR in our region despite a high rate of antibiotic consumption in our country. These results highlight the need for implementation of real-time AR surveillance systems which use factual data.

## INTRODUCTION

Even before the use of antibiotics, the phenomenon of antibiotic resistance existed^1^. It has evolved in several ecosystems under the influence of the use of antibiotics in animals (farm production) and in humans (healthcare-associated infections)^2^. It is difficult to estimate the burden of resistance to multiple antibiotics due to the use of multiple definitions and the lack of empirical data^3^. Many reports estimating mortality and morbidity due to antibiotic resistance have been published in recent years^4^. The most recent study from Cassini et al. estimated that antibiotic resistance was responsible for 33,110 deaths per year in Europe^5^. However, this report uses mathematical models that does not represent factually resistance to several classes of antibiotics in a given bacterial species or the real cost of antibiotic resistance on mortality^3,6–8^. These mathematical models, depending on the prevalence of antibiotic resistance in some countries, also use incidence ratios of a given infection to an invasive infection, and then mortality ratios, all from different literature reviews that could not be accurate to definitively estimate the number of extra deaths due to multidrug-resistance (MDR)^9^ (https://reflectionsipc.com/2018/11/07/amr-deaths-in-europe/). Indeed, as it is not matter of facts to acknowledge these conclusions you need to agree with the deductive method that being a question of opinion. Therefore, these statistics are subject to controversies reporting facts may be subject to discussion on their generalization but not on their reality.

From an epidemiological point of view, the classification of bacteria into MDR, extensively drug-resistant (XDR) and Pandrug resistant (PDR) has an interest, but is not clinically relevant since many other antibiotics could be tested and used to treat such infections if needed^10,11^. Recently, some reports have suggested other definitions based on the use of first-line antibiotics in patients that are more suitable for clinicians^4,12,13^. Kadri et al. suggested a new definition as “difficult-to-treat” (DTR) bacterial infections i.e infections due to bacteria that are *in vitro* resistant to all antibiotics tested in 3 classes of first-line antibiotics (ß-lactams, carbapenems and fluoroquinolones)^4^. According to this definition, they have reported only 1% of Gram-negative bacteria (GNB) classified as DTR in a large series of bacterial isolates from 173 hospitals in the USA over a 3 years period. In all cases, a therapeutic alternative was possible^4^.

Recent retrospective studies conducted in our laboratory hospital demonstrated an overall stability or a decrease in antibiotic resistance over the last decade^13^ with only one patient who died with a DTR bacterial infection^3,14^. Similarly, a recent survey conducted in 251 intensive care units (ICU) in France estimated about 45 deaths attributable to antibiotic resistance without alternative treatment over a 10-years period contradicting the prediction based on mathematical models^6^.

This disparity between reality and the myth of antibiotic resistance could only be resolved by implementing efficient antibiotic resistance surveillance systems that observe in real or near-real-time the results of susceptibility testing in deceased and survivors patients, as it is already the case in our institution^6^.

Therefore, at the Marseille University Hospital Institute, using the monitoring systems implemented BALYSES (Bacterial real-time Laboratory-based Surveillance)^15^, MARSS (Marseille Antibiotic Resistance Surveillance System) and PACASurvE (PACA Surveillance Epidemiologic System)^16^, we monitor weekly the results of all strains isolated in the 4 University Hospitals of Marseille (Assistance Publique Hôpitaux de Marseille) and in laboratories of the region which participate to this surveillance system for which antibiotic susceptibility testing (AST) results are available^6,15,16^.

Based on these different data sources, we retrospectively analyzed AST data from this network over a period of 5 years, focusing on the15 most frequently isolated bacteria that are clinically relevant in human diseases. We evaluated the resistance rate of bacteria to predefined key antibiotics and the evolution of this rate over time. Finally, we determined the number of DTR bacteria.

## MATERIAL AND METHODS

### Clinical settings

We conducted a retrospective study on AST of bacteria isolated in the Provence-Alpes-Côte d’Azur (PACA) region. This region of southeastern France has a population of 5,059,473 inhabitants and an area of 31,400 km2 (https://www.insee.fr/fr/statistiques/1893198). The data analyzed were collected from PACASurvE ^16^ and BALYSES ^15^ from January 2014 to January 2019. The data analyzed with BALYSES are those routinely produced by the AP-HM clinical microbiology laboratory, while PACASurvE analyzes are the data routinely produced by 303 different clinical microbiology laboratories, including 16 public hospital laboratories and 10 private laboratory groups from the PACA French region. In this study, we analyzed data from 267 laboratories for which AST data were available.

### Methodological steps

The flowchart (Figure 1) describes the main methodological steps followed in this study for the 15 most common bacteria isolated in our clinical microbiology laboratories, including *Escherichia coli, Klebsiella pneumoniae, Enterobacter cloacae, Kelsbiella aerogenes, Proteus mirabilis, Acinetobacter baumannii, Pseudomonas aeruginosa, Morganella morganii, Enterococcus faecium, Enterococcus faecalis, Staphylococcus aureus, Staphylococcus epidermidis, Streptococcus agalactiae, Serratia marcescens* and *Klebsiella oxytoca* ^15^. To harmonize results between the different laboratories of the PACA region, key antibiotics were selected as shown in Table 1. All strains with intermediate resistance were considered resistant for statistical analysis. AST was performed according to the EUCAST (European Committee on Antimicrobial Susceptibility Testing) recommendations.

**Table 1:**
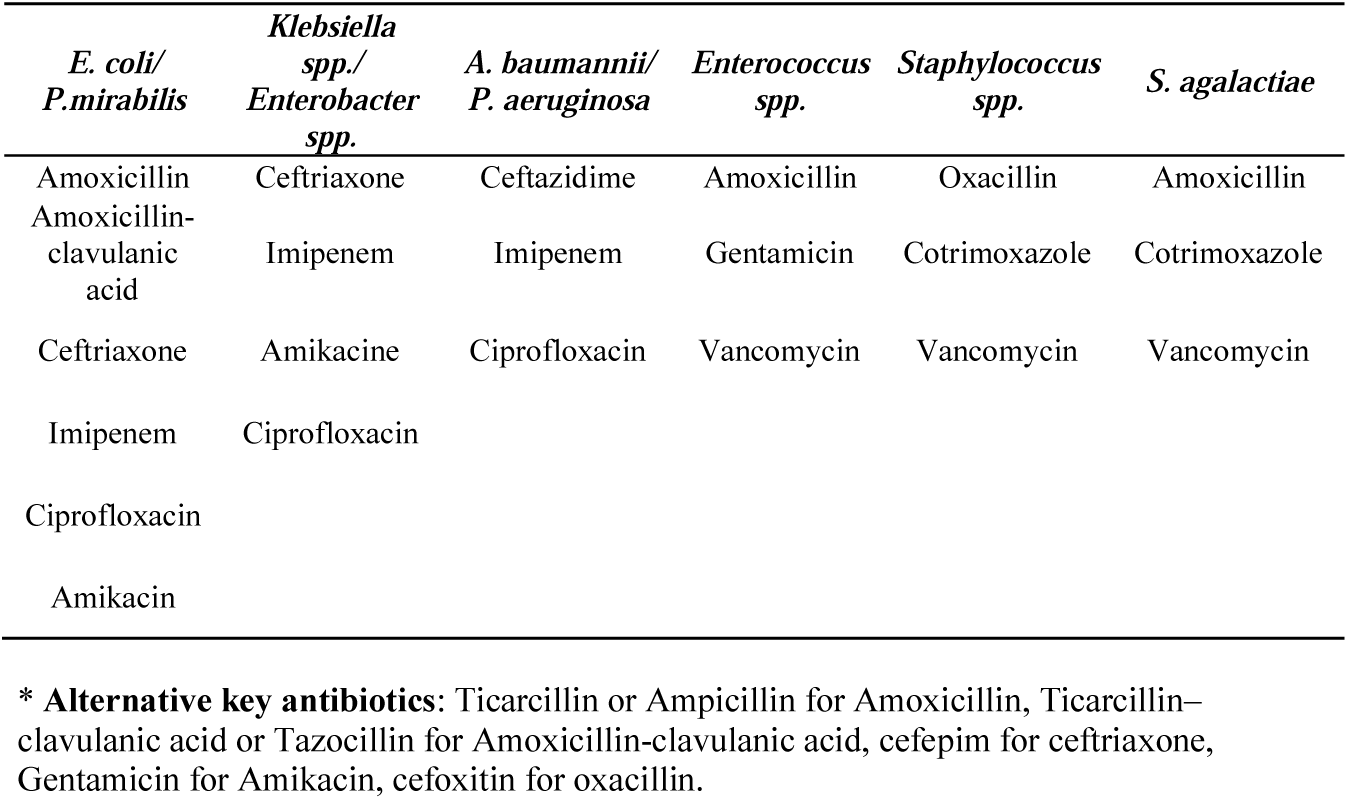
List of key antibiotics* chosen in this study.

| *E. coli/*<br>*P.mirabilis* | *Klebsiella*<br>*spp./*<br>*Enterobacter*<br>*spp.* | *A. baumannii/*<br>*P. aeruginosa* | *Enterococcus*<br>*spp.* | *Staphylococcus*<br>*spp.* | *S. agalactiae* |
| --- | --- | --- | --- | --- | --- |
| Amoxicillin | Ceftriaxone | Ceftazidime | Amoxicillin | Oxacillin | Amoxicillin |
| Amoxicillin-clavulanic<br>acid | Imipenem | Imipenem | Gentamicin | Cotrimoxazole | Cotrimoxazole |
| Ceftriaxone | Amikacine | Ciprofloxacin | Vancomycin | Vancomycin | Vancomycin |
| Imipenem | Ciprofloxacin |  |  |  |  |
| Ciprofloxacin |  |  |  |  |  |
| Amikacin |  |  |  |  |  |

\* **Alternative key antibiotics:** Ticarcillin or Ampicillin for Amoxicillin, Ticarcillin– clavulanic acid or Tazocillin for Amoxicillin-clavulanic acid, cefepim for ceftriaxone, Gentamicin for Amikacin, cefoxitin for oxacillin.

**Figure 1.**
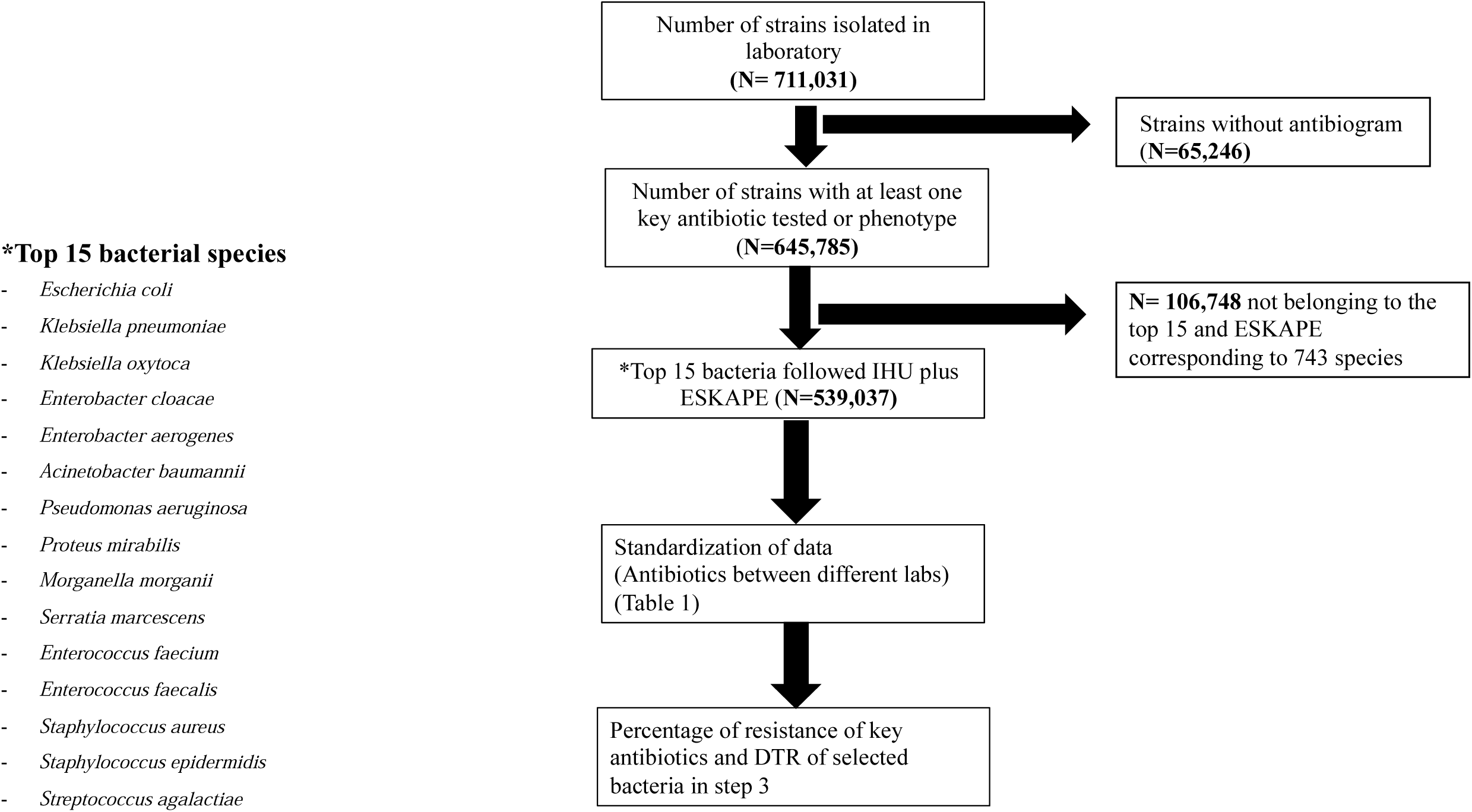
Flow chart of raw antibiogram data available January 2014-February 2019)

We also look for DTR bacteria following definition previously established by Kadri *et al*. for GNB ^4^. Briefly, a GNB was considered as DTR if it was resistant to all cephalosporins and penicillin’s+ inhibitor, all carbapenems and all fluoroquinolones. For Gram positive bacteria, the DTR definition were as follows: *Staphylococcus spp*. isolate was considered DTR if it was resistant to methicillin, gentamicin and vancomycin, whereas *Enterococcus spp*. should be resistant to at least amoxicillin, gentamicin and vancomycin to be classified as DTR. All criteria to define DTR bacteria are provided in Table 2. We then established resistance profiles for other antibiotics tested, in order to define therapeutic alternative for these bacteria.

**Table 2.** Phenotypic Definitions of Difficult-to-Treat Resistance.

| Gram negative bacteria | β-lactam | Extended-spectrum cephalosporin | Carbapenems | Fluoroquinolones |
| --- | --- | --- | --- | --- |
| *Escherichia coli*<br>*Klebsiella spp*<br>*Enterobacter spp*<br>*Proteus mirabilis*<br>*Serratia marcescens*<br>*Morganella morganii* | Aztreonam,<br>Piperacillin-tazobactam | Cefepime, Ceftriaxone,<br>Cefotaxime | Imipenem, Meropenem<br>Doripenem Ertapenem | Ciprofloxacin, Levofloxacin,<br>Moxifloxacin |
| *Acinetobacter baumannii* | Piperacillin-tazobactam | Ceftazidime, Cefepime | Imipenem, Meropenem<br>Doripenem | Ciprofloxacin, Levofloxacin,<br>Moxifloxacin |
| *Pseudomonas aeruginosa* | Aztreonam,<br>Piperacillin-tazobactam | Ceftazidime, Cefepime | Imipenem, Meropenem<br>Doripenem | Ciprofloxacin, Levofloxacin |
| Gram positive bacteria | β-lactam | Glycopeptides | | Aminosides |
| *Enterococcus faecium*<br>*Enterococcus faecalis* | Amoxicillin | Vancomycin, Teicoplanin |  | Gentamicin, Tobramycin |
| *Staphylococcus spp* | Oxacilline | Vancomycin, Teicoplanin |  | Gentamicin, Tobramycin |
| *Streptococcus agalactiae* | Amoxicillin | Vancomycin, Teicoplanin |  | Gentamicin |

### Statistical analysis

Resistance to key antibiotics ratios were determined for the complete data set. These ratios were compared with the Mann-Whitney U test, the χ2 test or the Fisher’s exact test and the Kendall test was used for correlation. For annual trend analysis, data for the year 2019 were not included in this study.

A p-value <0.05 was considered statistically significant. The R software (The R Project, Auckland, New Zealand) has been used to analyze the data.

## RESULTS

A total of 711,031 strains were isolated in all laboratories of the PACASurvE network, including 539,037 that belong to the 15 most common bacteria plus ESKAPE and had at least one key antibiotic tested (Figure 1). These data were recovered from 267 laboratories grouped in 6 hospitals laboratories and 7 groups of private laboratories. Urines (292,489; 54.26%) were the most prevalent samples followed by blood cultures (61,103; 11.34%), deeper samples (56,886; 10.55%), respiratory samples (46,966; 8.71%), skin samples (31,924; 5.92%), genital samples (27,562; 5.11%), ears-nose-throat samples (17,008; 3.16%), stools (4,611; 0.56%) and cerebrospinal fluid (488; 0.09%) (Figure 2. A). The fifteen most common bacterial species represented 539,037 (83.5%) isolates: *E. coli* accounted for 38% (246,353) of the isolates, followed by *S. aureus* (65,023; 10%), *K. pneumoniae* (49,733; 8%) and *E. faecalis* (36,857; 6%) (Table S1, Figure 2. B). These strains were isolated from 345,741 patients with most women (51.5%) and an average age of 54.6 years old.

**Figure 2:**
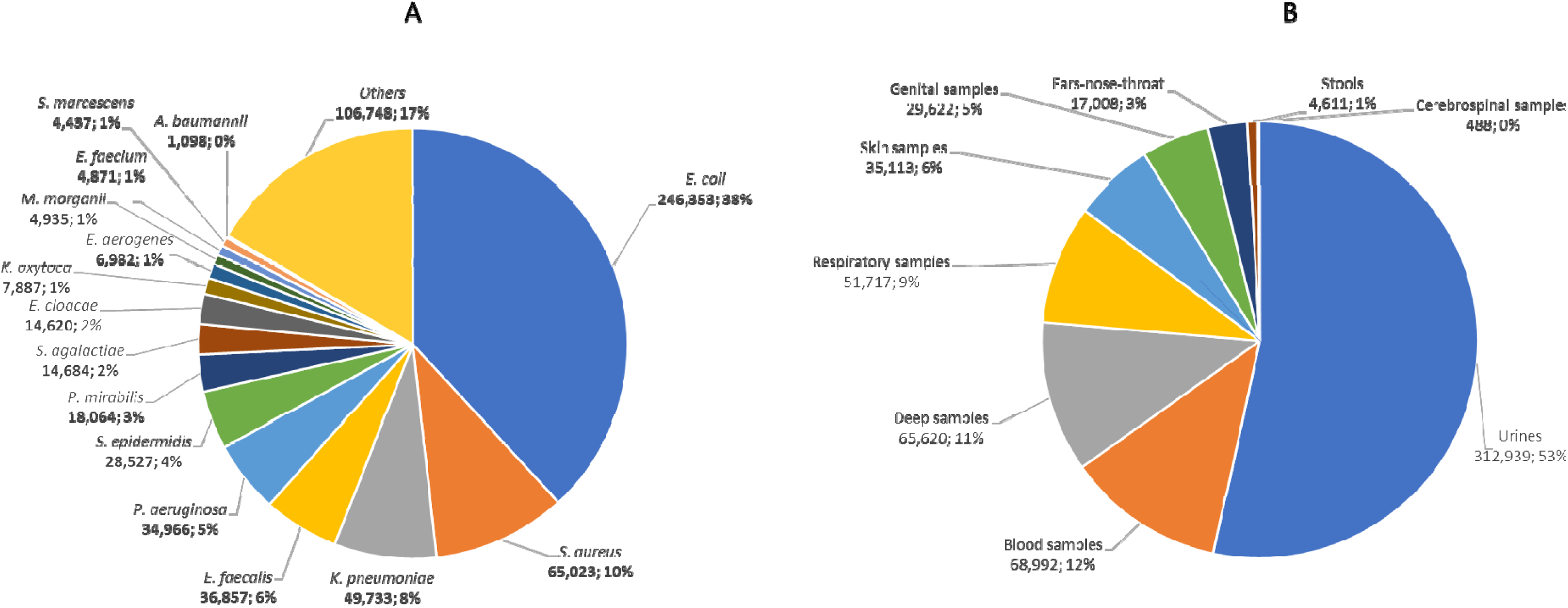
**A**. Presentation of the top 15 most frequently isolated bacteria in our surveillance systems (BALYSES and PACASurvE) between 2014 to 2019. **B**. The different types of samples (for only the laboratories that notify them).

### Proportions of resistant isolates

The proportion of resistant isolates for key antibiotics are shown in Table 3.

**Table 3.** Resistance rate to key antibiotics for the 15 most frequently isolated bacteria in Provence-Alpes-Côte d’Azur region.

|  | Number of infections | Rate % |
| --- | --- | --- |
| *Escherichia coli* |  |  |
| Amoxicillin-resistance | 140,588 | 50.4 |
| Amoxicillin-clavulanic acid resistance | 59,909 | 25.5 |
| 3GC-resistance | 25,650 | 9.5 |
| Ciprofloxacin resistance | 37,549 | 14.9 |
| Carbapenem resistance | 202 | 0.2 |
| Amikacin resistance | 6,126 | 2.5 |
| *Proteus mirabilis* |  |  |
| Amoxicillin resistance | 8,272 | 40.7 |
| Amoxicillin-clavulanic acid resistance | 1,811 | 10.2 |
| 3GC-resistance | 353 | 1.9 |
| Ciprofloxacin resistance | 2,248 | 11.9 |
| Carbapenem resistance | 195 | 2.4 |
| Amikacin resistance | 406 | 2.4 |
| *Klebsiella pneumoniae* |  |  |
| 3GC-resistance | 12,404 | 25.8 |
| Carbapenem resistance | 600 | 1.9 |
| Amikacin resistance | 2,326 | 5.2 |
| *Klebsiella oxytoca* |  |  |
| 3GC-resistance | 732 | 10.1 |
| Carbapenem resistance | 30 | 0.5 |
| Amikacin resistance | 323 | 4.7 |
| *Klebsiella aerogenes* |  |  |
| 3GC-resistance | 811 | 16.9 |
| Carbapenem resistance | 76 | 1.6 |
| Amikacin resistance | 248 | 4.1 |
| *Enterobacter cloacae* |  |  |
| 3GC-resistance | 3,180 | 36.1 |
| Carbapenem resistance | 179 | 1.2 |
| Amikacin resistance | 797 | 5.6 |
| *Acinetobacter baumannii* |  |  |
| 3GC-resistance | 508 | 50.0 |
| Carbapenem resistance | 225 | 20.3 |
| Ciprofloxacin resistance | 456 | 47.0 |
| Amikacin resistance | 159 | 25.6 |
| *Pseudomonas aeruginosa* |  |  |
| 3GC-resistance | 4,673 | 13.2 |
| Carbapenem resistance | 7,191 | 19.6 |
| Ciprofloxacin resistance | 5,924 | 21.2 |
| Amikacin resistance | 2,544 | 9.2 |
| *Enterococcus faecalis* |  |  |
| Amoxicillin resistance | 84 | 0.3 |
| Gentamicin resistance | 7,400 | 32.1 |
| Vancomycin resistance | 34 | 0.1 |
| *Enterococcus faecium* |  |  |
| Amoxicillin resistance | 3,168 | 82.1 |
| Gentamicin resistance | 1,856 | 54.1 |
| Vancomycin resistance | 100 | 2.9 |
| *Staphylococcus aureus* |  |  |
| Methicillin resistance | 9,295 | 16.9 |
| Cotrimoxazole resistance | 564 | 1.1 |
| Vancomycin resistance | 0 | 0.0 |
| *Streptococcus agalactiae* |  |  |
| Penicillin resistance | 7 | 0.07 |
| Cotrimoxazole resistance | 184 | 3.1 |
| Vancomycin resistance | 0 | 0.0 |

(Number of strains resistant/Number of AST\*\*)

Globally, we observed a significant decrease in amikacin resistance from 2014 to 2018 (792/16,733; 4.7% to 1,105/80,977; 1.36%, p=0.04) for *E. coli, K. pneumoniae* (363/3,963; 9.16% to 448/13,804; 3.25%, p=0.004), *P. mirabilis* (76/1,390; 5.47% to 55/5,435; 1.01%, p=0.01) and *K. oxytoca* (63/651; 9.68% to 77/2,055; 3.75%, p=0.006) (Figure 3 and Figure S1, Table S2). We also observed a significant decrease in imipenem resistance for *A. baumannii* from 2014 to 2018 (66/229; 28.82% to 23/185; 12.43%; p=0.005). However, we noticed a significant increase in ceftriaxone resistance in *E. aerogenes* (38/389; 9.77% to 307/1,470; 20.88%; p=0.001), *K. oxytoca* (38/389; 9.77 % to 307/1,470; 20.88 %, p=0.001) and *E. cloacae* (187/861; 21.72% to 1,305/2,746; 47.52%; p=0.004) whereas it remains stable in *E. coli* (2,046/21,067; 9.71 % to 7,551/84,657; 8.92 %, p=0.88), *K. pneumoniae* (1,178/4,400; 26.77% to 3,413/14,413; 23.68%, p=0.89), *P. mirabilis* (41/1,880; 2.18 % to 88/5,749; 1.53%, p=0.87). For *E. faecalis* we observed a significant increase in gentamicin resistance (356/2,774; 12.83% to 4,215/6,446; 65.39%, p=0.004). We did not observe any significant increase or decrease in resistance to key antibiotics for the other bacterial species studied.

**Figure 3.**
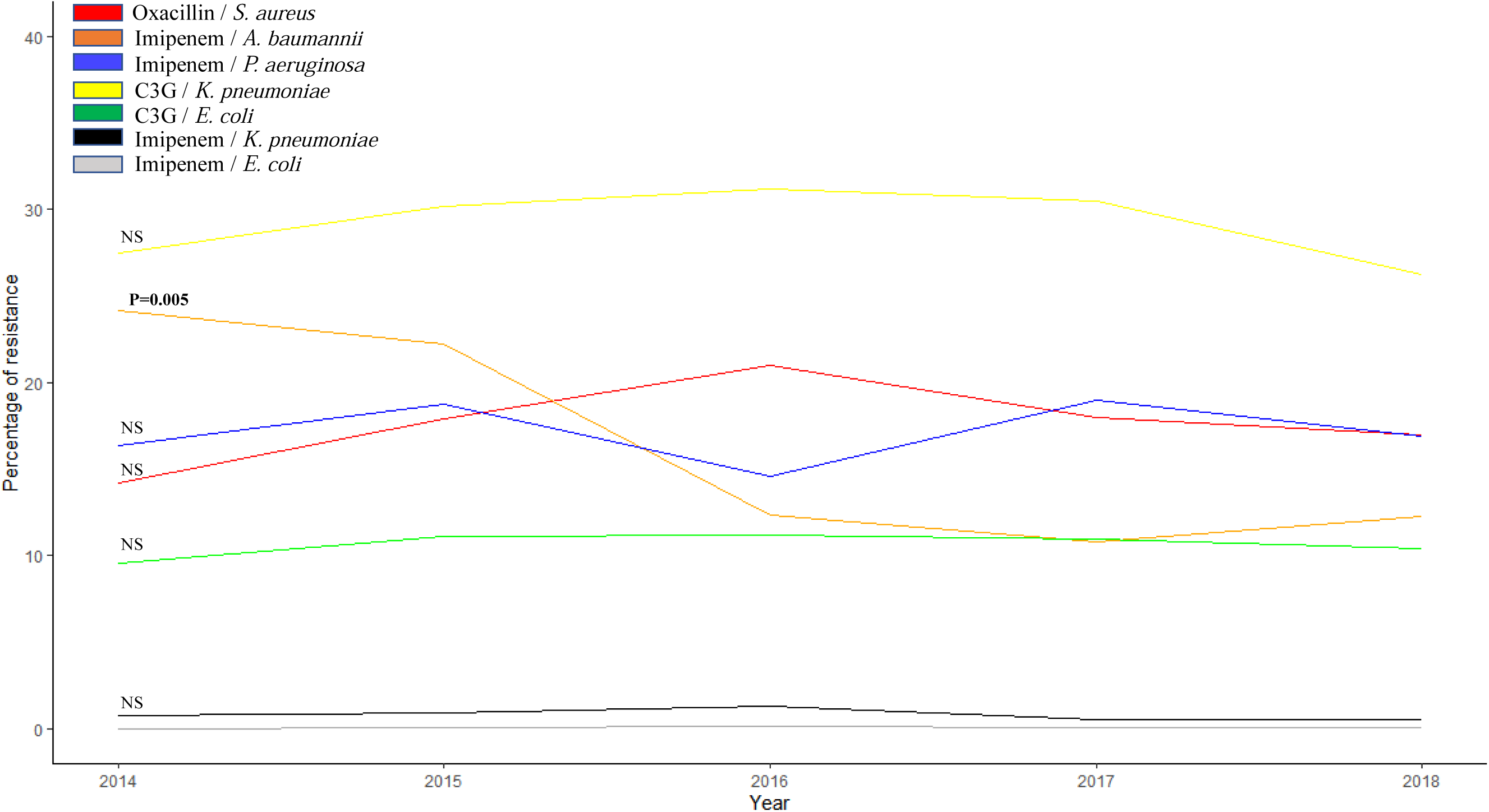
Evolution of resistance percentage of key antibiotics in bacterial species isolated from January 2014 to December 2018.

### Percentage of bacteria classified as Difficult-to-threat

Of the 539,037 bacterial strains belonging to the top 15 plus ESKAPE analyzed in this study, 1,604 strains (0.3%) carried a DTR phenotype (Table 4). These isolates were mostly GNB, and we identified 11 Gram positive bacteria with a DTR phenotype (11/1,604;0.68%). Among GNB, *A. baumannii* was the most prevalent bacterium carrying a DTR phenotype (175; 15.9%), followed by *P. aeruginosa* (902; 2.6%) and *K. pneumoniae* (372; 0.7%). However, we observed an overall evolution of non-significant DTR during our study period (Figure S2). The overall rate of DTR isolates in GNB was significantly lower (0.44% vs 1%, p<10^−5^) than that observed in Kadri *et al*. study ^4^. In both studies, *A. baumannii* was the most prevalent bacterium frequently considered as difficult-to-treat (15.33% and 18.3%, p=0.24), followed by *P. aeruginosa* (2.63% and 2.3% p=0.21) and *Klebsiella spp*. (0.66% and 1.7%, p**<** 10^−5^).

**Table 4.** Prevalence of strains carrying a Difficult-to-treat Resistance (DTR) phenotype isolated in Provence-Alpes-Côte d’Azur from January 2014 to February 2019 and comparison with the study of Kadri et al.

| Bacterial strains | Number of DTR<br>(Number of strains) | Rate (%) | 173 hospitals in USA (Kadri<br>et al. 2018) | P_value |
| --- | --- | --- | --- | --- |
| *Escherichia coli* | 85 (246, 353) | 0.03 | 12/28,640 (0.04) | 0.64 |
| *Klebsiella spp. (K. pneumoniae and K. oxytoca)* | 383 (57,620) | 0.66 | 155/9,168 (1.7) | $< 10^{-5}$ |
| *Enterobacter spp. (K. aerogenes\* and E. cloacae)* | 39 (21,602) | 0.18 | 20/3,221 (0.6) | $< 10^{-5}$ |
| *Proteus mirabilis* | 1 (18,064) | 0.006 | NA |  |
| *Serratia marcescens* | 3 (4,437) | 0.07 | NA |  |
| *Morganella morganii* | 5(4,935) | 0.1 | NA |  |
| *Pseudomonas aeruginosa* | 902 (34,966) | 2.6 | 101/4,493 (2.3) | 0.21 |
| *Acinetobacter baumannii* | 175 (1,098) | 15.9 | 183/999 (18.3) | 0.24 |
| *Enterococcus faecalis* | 3 (36,857) | 0.008 | NA |  |
| *Enterococcus faecium* | 5 (4,871) | 0.10 | NA |  |
| *Staphylococcus aureus* | 0 (65,023) | 0 | NA |  |
| *Staphylococcus epidermidis* | 3 (28,527) | 0.01 | NA |  |
| *Streptococcus agalactiae* | 0 (14,684) | 0 | NA |  |
| Total (comparison with Kadri<br>et al.) | **1,584 (361,639)** | **0.44** | **1.01 (471/46,521)** | $< 10^{-5}$ |
| **Total** | **1, 604 (539,037)** | **0.3** | NA |  |

\*Previously *Enterobacter aerogenes*, having resistance similar to *Enterobacter spp* species.

## DISCUSSION

Nowadays, mathematical models based on predictions are of major importance in public health decision-making. While they have an interest in trying to assess what might happen in the future, they essentially require confirmation or refutation by factual data that cannot be contested. Factual data sometimes have the disadvantage of being different from one laboratory to another, but the harmonization of microbiological practices, through EUCAST or CLSI, tends to correct this discrepancy as it was the case in our study. Moreover, we were not always able to know if some isolates were from the same patient, but such bias was corrected by the mass of data analyzed. However, as our data came from a large but limited number of laboratories in the PACA region, they cannot be extrapolated to the entire region, nor to France or Europe. Thus, in our study, we did not observe any significant increase or decrease in resistance to key antibiotics, including oxacillin resistance in *S. aureus*, carbapenem resistance in enterobacteria and *P. aeruginosa*, and 3^rd^ generation cephalosporin in *E. coli* and *K. pneumoniae*.

To our knowledge, this study is the world’s largest series analyzing data on antibiotic resistance over a 5-year period, which allows to appreciate our local epidemiology on antibiotic resistance and to draw reliable conclusions on global trends. In literature, we found only three major world series that tested more than 100,000 isolates ^17–19^ (Table 5), but they were limited to *E. coli* strains, unlike our study which focused on 15 bacterial species and their key antibiotics and were not recent studies (Table 1). Two of these studies occurred on a ten years period. The first one took place in Austria from January 1998 to December 2013, focused on 135,878 *E. coli* strains and showed a significant increase in amoxicillin, 3^rd^ generation cephalosporin, ciprofloxacin and cotrimoxazole resistance^17^. The second one took place from January 2009 to October 2013 in Spain on 141,583 *E. coli*^18^. They showed only a change in resistance to amoxicillin and clavulanic acid that has increased over the years 20. Finally, the third study conducted in the United Kingdom from January 1996 to December 2016 included 228,374 strains of *E. coli* over a 4-year period but did not analyze the level of resistance over the years^19^.

**Table 5.**
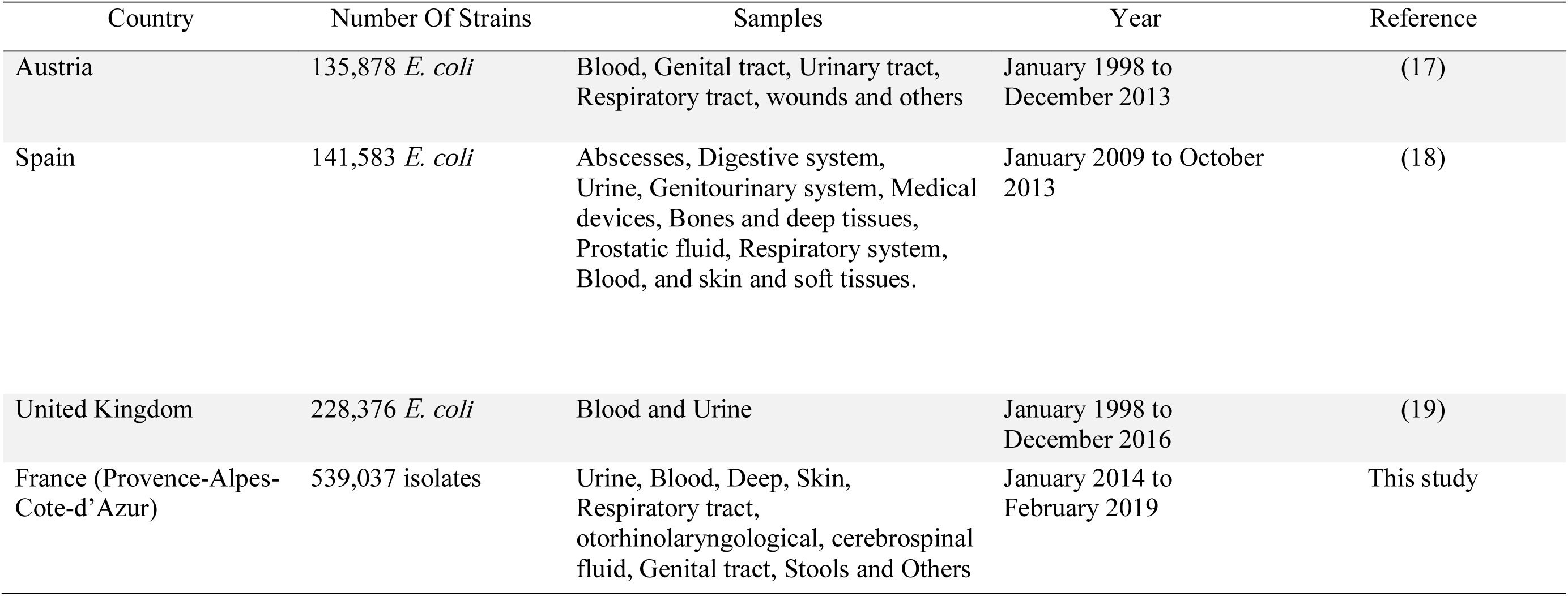
The world’s largest series on the study on antibiotic resistance.

The number of strains classified as DTR in our study was very low (1,604/539.037; 0.3%) and constitutes a very rare event (<1%). This rate was lower than that observed in the study conducted by Kadri *et al*. from hospitals in the USA. This finding may be explained by our recruitment: we include both hospital and community-acquired isolates and we did not focus only on bacteremia and include all type of samples.

Thus, our study did not show a worrying increase in resistance to key antibiotics in our region over a 5-year period. Interestingly, despite the fact that France is considered the largest consumer of antibiotics in Europe^20^, and the PACA region is above the national average (31.6 DDJ/1000H/J) according to the national survey (https://ansm.sante.fr/var/ansm_site/storage/original/application/188a6b5cf9cde90848ae9e3419bc3d3f.pdf) (Figure 4), we did not observe an increase in antibiotic resistance in our study. Predictive models are dependent on the belief in these models and cannot replace the facts that remain at the end. We believe that “real time” surveillance systems are the only ones capable of detecting abnormal or emerging bacterial resistance in the community and/or in hospitals. This real-time monitoring of antibiotic resistance is mandatory because the main factor associated with mortality and antibiotic resistance is an inappropriate initial antibiotic treatment and not the resistance to a single antibiotic^6^. As mentioned above, the relevance of surveillance systems depends on the number of data collected. Nowadays, data collection is becoming increasingly simple with technological advances and the implementation of automatic computerized systems. However, the General Data Protection Regulations legislation limit access to this data, even anonymized. These data are the foundation of these monitoring systems and are essential for research and public health. Thus, they are critical to adapt empirical therapeutic strategies according to local epidemiology because antibiotic resistance is a complex phenomenon that is unpredictable^21,22^. Moreover, only these observations rather than prediction of a virtual future should be taken in account to make public health decision. There is an urgent need to find a balance that can guarantee the protection of patients’ data, without limiting scientific research. Finally, in order to avoid the current fear of antibiotic resistance worldwide, we believe that it is urgent to set up sustainable real registers of deaths and antibiotic resistance instead of mathematical models.

**Figure 4.**
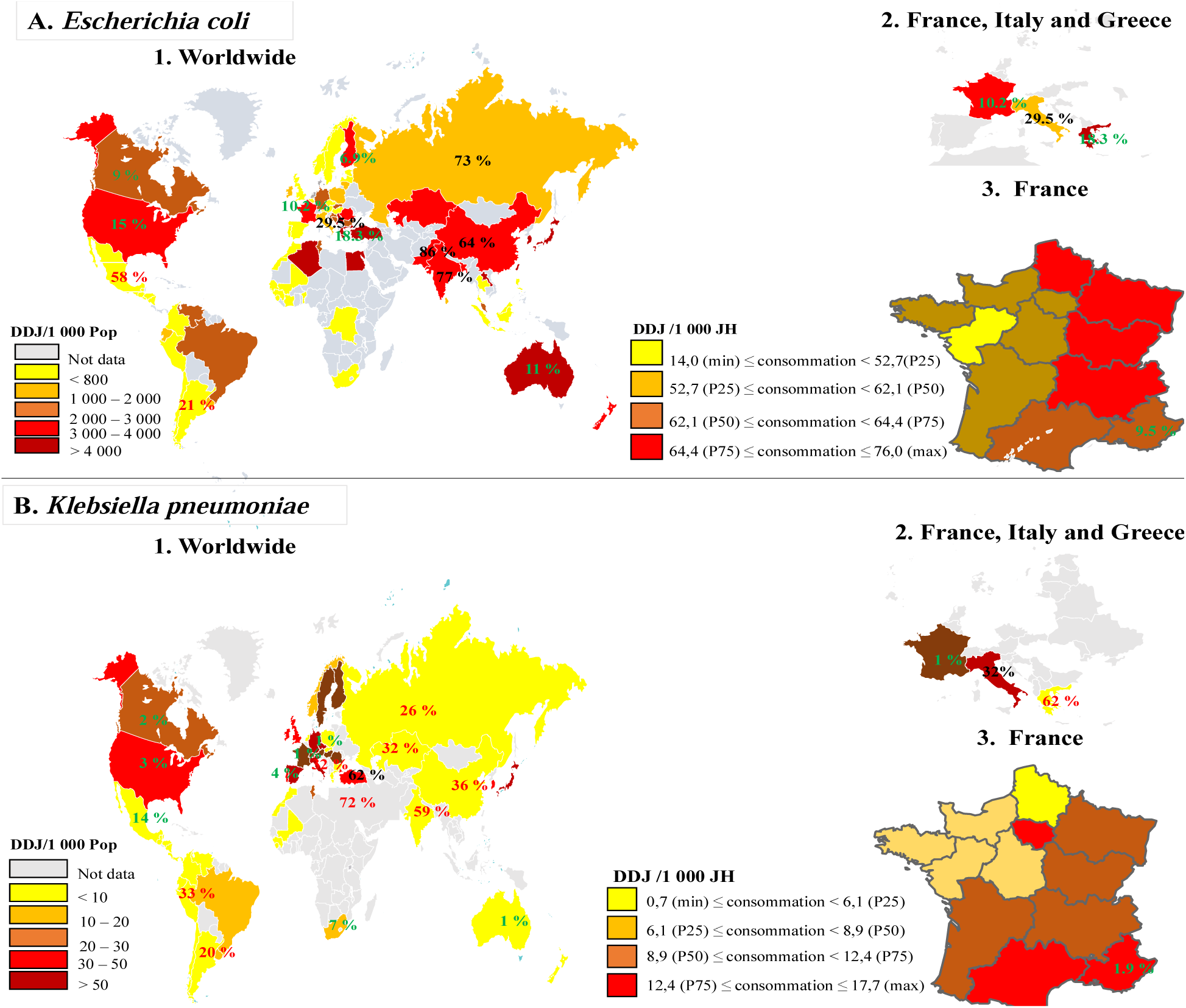
Third Cephalosporin generation consumption and resistance (%) worldwide.

## Data Availability

All data are available in the supplementary files

## Funding information

This work was supported by the French Government under the «Investissements d’avenir» (Investments for the Future) program managed by the Agence Nationale de la Recherche (ANR, fr: National Agency for Research), (reference: Méditerranée Infection 10-IAHU-03)

## Acknowledgment

We want to thank CookieTrad for English correction.

## Transparency declaration

The authors declare that they have no competing interests

## Author’s contribution

OOD, SAB, DR, JMR designed the study, drafted and revised the manuscript.

GD, PHal, SC, BC, SJ, AF, PSe, FP, JPC, RR, CM, JCA, SB, JC, ED, SM, PYL, PR, PB, CB, PSt, JPA, GG, PHan, PC performed, retrieved and analyzed microbiology analyses. OOD, SAB, HC and JMR performed statistical analyses

All authors have read and approved the final manuscript.

## Supplementary data: 4

Table S1: Presentation of the different bacterial species isolated from our surveillance system from January 2014 to February 2019 (N=711,031).

Table S2: Trend in antibiotic resistance of the species studied for the full years 2014 to 2018.

Figure S1. Evolution of resistance percentage of key antibiotics in bacterial species isolated by the different laboratories included in our study from January 2014 to December 2018.

**A**. *E. coli;* **B**. *P. mirabilis*; **C**. *E. cloacae***; D**. *E. aerogenes***; E**. *K. oxytoca***; F**. *S. marcescens***; G**. *K. pneumoniae;* ***H***. *M. morganii;* **I**. *E. faecalis;* **J**. *E. faecium;* **K**. *S. epidermidis;* **L**. *S. agalactiae*

Figure S2. Evolution of the number of strains carrying a DTR phenotype between 2014 and 2018.

## Notes

### Competing Interest Statement

The authors have declared no competing interest.

